# Human *Paenibacillus* Infections: A Systematic Review with Comparison of Adult and Infant Cases

**DOI:** 10.1101/2023.09.19.23295794

**Authors:** Danielle Smith, Kristen Bastug, Kathy Burgoine, James R. Broach, Christine Hehnly, Sarah U. Morton, Marwan Osman, Steven J. Schiff, Jessica E. Ericson

## Abstract

Neonatal infections due to *Paenibacillus species* have increasingly been reported over the last few years. We performed a structured literature review of human *Paenibacillus* infections in infants and adults to compare the epidemiology of infections between these distinct patient populations. Thirty-nine reports describing 176 infections met our inclusion criteria and were included. There were 37 *Paenibacillus* infections occurring in adults caused by 23 species. The clinical presentations of infections were quite variable. In contrast, infections in infants were caused by only 3 species: *P. thiaminolyticus* (112/139, 80%), *P. alvei* (2/139, 1%) and *P. dendritiformis* (2/139, 1%). All of the infants with *Paenibacillus* infection presented with a sepsis syndrome or meningitis, often complicated by extensive cerebral destruction and hydrocephalus. Outcomes were commonly poor with 17% (24/139) mortality. Cystic encephalomalacia due to brain destruction was common in both Ugandan and American cases and 92/139 (66%) required surgical management of hydrocephalus following their infection. *Paenibacillus* infections are likely underappreciated in infants and effective treatments are urgently needed.

---

Neonatal sepsis is a rare but devastating disease affecting infants worldwide.^1^ Over 2 million deaths occur each year and even for survivors of neonatal sepsis, sequelae include neurodevelopmental delays, hearing impairment, seizure disorders and hydrocephalus.^2^ The etiology of neonatal sepsis often remains unknown especially in low-resource settings.^3^ Bacterial culture of the blood and spinal fluid sometimes identifies the causative pathogen but is limited by: 1) access to laboratory facilities that can perform cultures; 2) reduced sensitivity of culture results due to community-based antibiotic administration prior to culture collection; or 3) inability to grow the causative pathogen due to the growth requirements of the organism. Technologic advances in molecular diagnostics have enabled improved detection and identification of microbial organisms in patient specimens throughout the world. Organisms previously considered environmental contaminants that are identified in patients who are critically ill could represent emerging pathogens, particularly in high-mortality illnesses such as neonatal sepsis.

In our long-term effort to identify the etiology of neonatal sepsis, meningitis and their much feared sequela of postinfectious hydrocephalus in Uganda, we identified *Paenibacillus* species to be both the most commonly identified genera in infants with postinfectious hydrocephalus^4^ and also present in 6% of neonates with clinical sepsis.^5^ Further, we have found that *Paenibacillus* species are variably detected by standard aerobic microbial culture, likely due to high requirements for thiamine in growth media.^6^ Since 2021, there have been increasing reports of paenibacilliosis in infants in the United States with severe sequelae occurring in most published cases. *Paenibacillus* may represent an unrecognized cause of global neonatal sepsis, meningitis and postinfectious hydrocephalus.

*Paenibacillus species’* are spore-forming facultative anaerobic Gram-variable rods. *Paenibacillus* was first described in the early 1990s as a separate genus from *Bacillus*, which was previously thought to encompass these bacteria.^7^ These organisms were historically believed to be primarily opportunistic, causing infection only in those who were immunocompromised or using illicit intravenous drugs.^8^ Subsequent case reports have suggested that a broader population, including infants, is at risk for severe infection. We performed a structured literature review of human *Paenibacillus* infections in infants and adults to compare the epidemiology of infections between these distinct patient populations.

## Methods

Using Pubmed, we used the search term “*Paenibacillus* infection” limited to human studies to identify manuscripts for review. The authors reviewed papers to confirm that they were about a human infection; those found to describe infections of a plant or insect were excluded as were those describing identification of the organism from environmental samples. The references of included papers were reviewed for additional papers meeting our inclusion criteria. Finally, because the *Paenibacillus* genus was reclassified from the *Bacillus* genus, we also searched for each *Paenibacillus* species using *Bacillus* in place of *Paenibacillus*. Patient-level data from full-text papers confirmed to be about human infection due to a *Paenibacillus* species were abstracted from each paper using a structured data collected instrument in Excel (Microsoft, Redmond, WA, USA).

## Results

Our search terms identified 84 papers for review (Figure 1). After review, 63 were excluded: 4 were not about *Paenibacillus*, 6 described *Paenibacillus* infections of plants or insects, 12 described isolation of *Paenibacillus* from the environment, 26 described *in vitro* studies of the bacteria, mostly related to its antimicrobial properties against other pathogens; 6 noted the presence of *Paenibacillus* as part of the human microbiome but were not about infection; 4 did not provide patient-level data; 2 discussed the role of *Paenibacillus* in food spoilage; 2 did not have full text versions of the manuscript available for review and were excluded. One additional paper included patients that were described in more detail elsewhere and was excluded. Twenty-one papers remained that described *Paenibacillus* infection in humans. Review of the references of these papers identified an additional 16 papers meeting our inclusion criteria and searches for each species using *Bacillus* as the genus identified an additional 2 cases. We ultimately included 176 cases of *Paenibacillus* infection, 37 in adults and 139 in infants. Cases in adults occurred sporadically over time; most cases in infants have been reported in the last 3 years (Figure 2).

**Figure 1.**
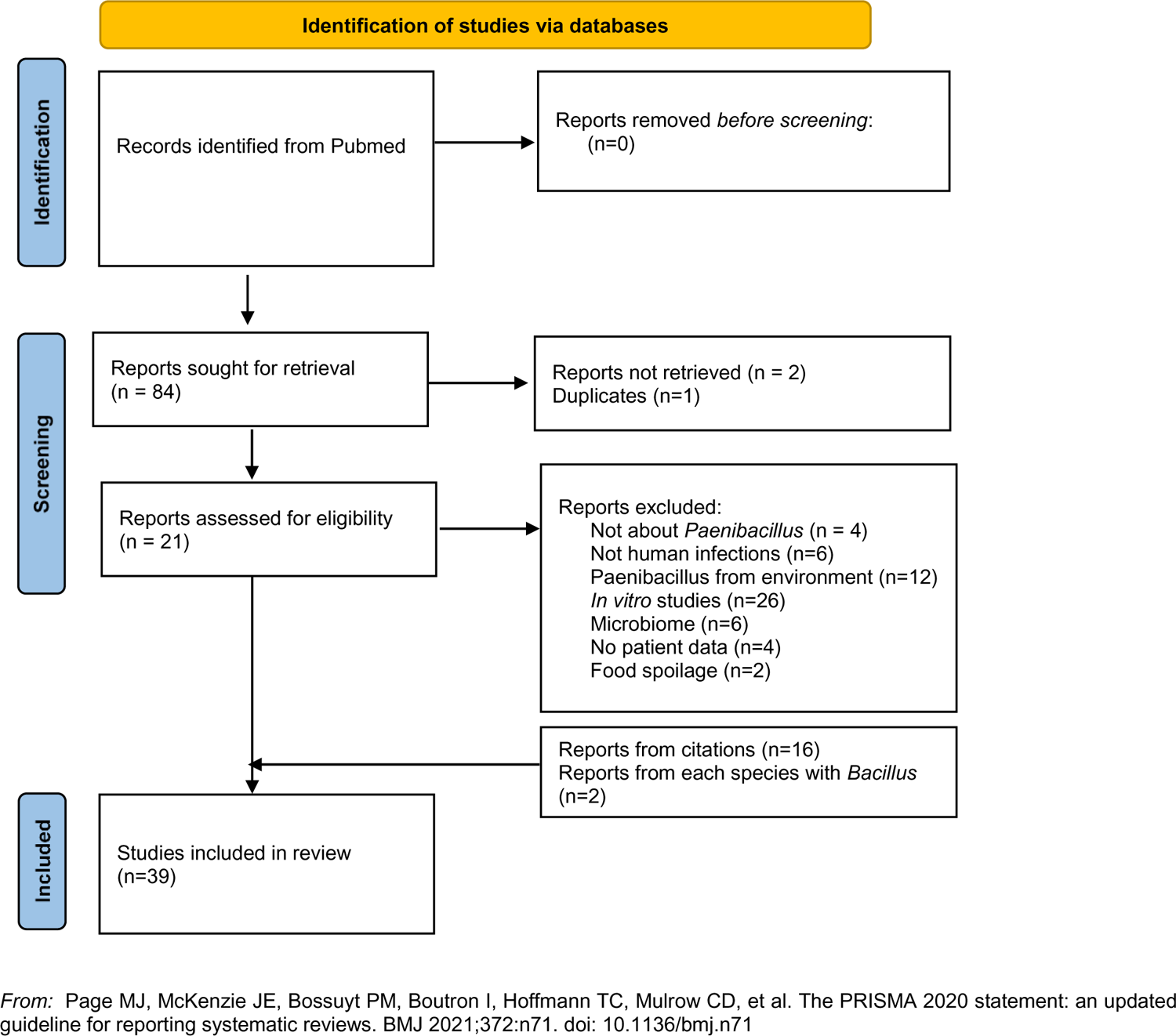

**Figure 2.**
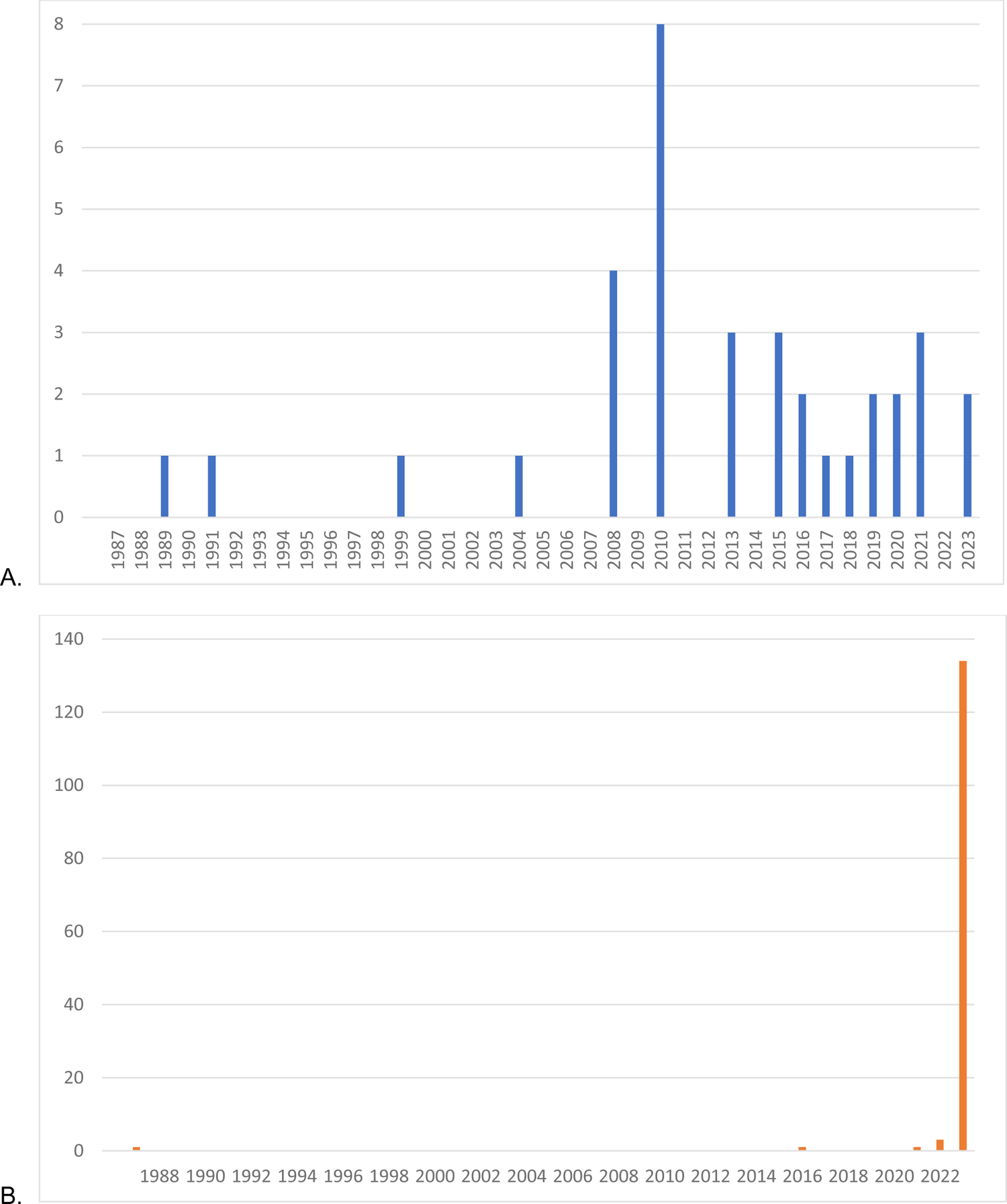
Annual published cases of infection due to *Paenibacillus species* in adults (A) and infants (B).

We identified 30 reports describing 37 *Paenibacillus* infections occurring in adults. Cases caused by 23 species were reported from 15 countries spanning 4 continents (Europe, n=21; Asia, n=8; North America, n=7, and South America, n=1) (Table 1). Surprisingly, there were no reports from Africa of Paenibacillus infections occurring in an adult; 27/37 (73%) were from developed countries, where laboratory technologies and other aspects of healthcare delivery are well established. The clinical presentations of the infections were also quite variable and included 16 bloodstream infections^8–17^, 3 of which occurred in the setting of endocarditis^14,15,17^ and 1 related to a dialysis catheter^13^; 8 wound infections^18–25^, 3 of which involved the bone^19,20,23^ and 1 involved a breast implant^18^; 3 central nervous system infections^26–28^ including 2 with a brain abscess^26,28^; 2 urinary tract infections^27,29^; 3 lung infections^30–32^; 2 joint infections^33,34^ one of which involved a prosthetic hip^34^; and 1 each of keratitis^35^, gingivitis^36^, superficial skin abscess^37^. There was a single death reported in a 55-year-old Korean man who developed *P. pasadensis* bacteremia following clipping of an intracranial aneurysm.^11^ His postoperative course was complicated by acute respiratory distress syndrome and he was intubated and mechanically ventilated for nearly two weeks before decompensating. Six of 6 blood cultures grew *P. pasadensis* and, due to the severity of his illness and complications that had already occurred prior to developing bacteremia, the focus of his care became palliative and he died a few days later. Three patients had a recurrence of their infection but were ultimately cured following repeat or long-term antibiotic therapy; the remaining 31 patients recovered. Interestingly, the 2 patients with brain abscess due to *P. lactis* and *P. macerans* fully recovered and had no neurologic sequelae after being treated with vancomycin, meropenem and metronidazole for an unknown duration and for 6 weeks, respectively.^26,28^

**Table 1.** Reported infections due to *Paenibacillus species* in adults.

| Year | Age, years | Organism | Clinical Illness | Comorbidity | Country | Treatment | Outcome |
| --- | --- | --- | --- | --- | --- | --- | --- |
| 1989<sup>34</sup> | 26 | *P. alvei* | prosthetic hip infection | sickle cell disease | USA | 6 weeks vancomycin, ceftriaxone | cured |
| 1991<sup>31</sup> | 62 | *P. alvei* | pneumonia, empyema | congestive heart failure | USA | penicillin | recurrence of pneumonia 1 year later, retreated and cured |
| 1999<sup>17</sup> | 57 | *P. popilliae* | Endocarditis | None | China | 6 weeks penicillin | cured |
| 2004<sup>9</sup> | 13 | *P. massiliensis* | bacteremia | leukemia | France | unknown | unknown |
| 2004<sup>9</sup> | 49 | *P. sanguinis* | bacteremia | oropharyngeal cancer | France | unknown | unknown |
| 2004<sup>9</sup> | 75 | *P. timonensis* | bacteremia |  | France | unknown | unknown |
| 2008<sup>10</sup> | 75 | *P. konsidensis* | bacteremia |  | Korea | unknown | unknown |
| 2008<sup>13</sup> | 80 | *P. thiaminolyticus* | bacteremia, dialysis catheter infection | ESRD, CHF, colon cancer | USA | 3 weeks vancomycin | unknown |
| 2008<sup>27</sup> | 54 | *P. provencesnsis* | positive CSF culture | Wippel's disease | France | none | unknown |
| 2008<sup>27</sup> | 36 | *P. urinalis* | urinary tract infection | IVDU | France | none | unknown |
| 2010<sup>8</sup> | 28 | *P. larvae* | bacteremia | IVDU | Germany | 14 days meropenem +<br>14 days penicillin | recurrence of BSI,<br>peritonitis then<br>cured |
| 2010<sup>8</sup> | 32 | *P. larvae* | bacteremia | IVDU | Germany | none | unknown |
| 2010<sup>8</sup> | 20 | *P. larvae* | bacteremia | IVDU | Germany | none | unknown |
| 2010<sup>8</sup> | 35 | *P. larvae* | bacteremia | IVDU | Germany | 7 days cefuroxime | unknown |
| 2010<sup>8</sup> | 27 | *P. larvae* | bacteremia | IVDU | Germany | 21 days imipenem | pulmonary<br>embolism then<br>cured |
| 2010<sup>32</sup> | 25 | *P. cineris* | pneumonia | cystic fibrosis | Brazil | 7 days amox-clav +<br>levofloxacin | cured |
| 2010<sup>30</sup> | 60 | *P. sputi* | pneumonia | NTM lung disease | Korea | unknown | unknown |
| 2010<sup>20</sup> | 79 | *P. pasadensis* | mediastinitis | myocardial infarction | Switzerland | 2 weeks<br>vancomycin/ciprofloxacin<br>+ 4 weeks<br>clindamycin/ciprofloxacin | cured |
| 2013<sup>15</sup> | 65 | *P.*<br>*glucanolyticus* | bacteremia,<br>endocarditis,<br>pacemaker<br>associated | diabetes, pacemaker | France | 6 weeks ceftriaxone | cured |
| 2013<sup>29</sup> | 60 | *P. alvei* | pyelonephritis |  | India | 7 days cefotaxime | cured |
| 2013<sup>21</sup> | 35 | *P. vulneris* | wound | unknown | Norway | not described | not described |
| 2015<sup>11</sup> | 55 | *P. pasadensis* | bacteremia | ARDS, intracranial<br>aneurysm clipping | Korea | teicoplanin, moxifloxacin,<br>meropenem | died |
| 2015<sup>28</sup> | 42 | *P. macerans* | brain abscess | Ischemic stroke,<br>craniectomy | USA | 6 weeks vancomycin,<br>cefepime, metronidazole | cured |
| 2015<sup>12</sup> | 36 | *P. amylolyticus* | bacteremia,<br>gingivitis | IVDU, trauma | USA | amox-clav | unknown |
| 2016<sup>18</sup> | 54 | *P. residui* | wound, breast<br>implant infection | breast cancer | Italy | amox-clav, surgical<br>drainage | cured |
| 2016<sup>23</sup> | 65 | *P. turicensis* | traumatic bone<br>infection | intramedullary nail | Laos | 6 months amoxicillin | cured |
| 2017<sup>14</sup> | 70 | *P. provencensis* | bacteremia,<br>endocarditis | myocardial infarction | England | 4 weeks<br>vancomycin/meropenem<br>+ 2 weeks daptomycin | cured |
| 2018<sup>33</sup> | 44 | *P. assamensis* | joint, septic knee | none | China | unknown | prolonged knee<br>symptoms |
| 2019<sup>36</sup> |  | *P. oralis* | gingivitis |  | Korea | unknown | unknown |
| 2019<sup>22</sup> | 66 | *P. macerans* | wound infection | diabetes, prostate<br>cancer | USA | tmp/smx | recurred so now on<br>suppressive<br>tmp/smx and doing<br>well |
| 2020<sup>37</sup> | 49 | *P. provencensis* | Skin abscess | none | Ireland | clindamycin, flucloxacillin<br>x 60 days | Cured |
| 2020<sup>24</sup> | 37 | *P. timonensis* | traumatic wound<br>infection | hypertension, NF1 | Spain | vancomycin x 10 days +<br>3 weeks tmp/smx | cured |
| 2021<sup>35</sup> |  | *P. glucanolyticus* | corneal abscess | IVDU | England | ocular gentamicin, po<br>doxycycline | infection resolved,<br>permanent vision<br>loss |
| 2021<sup>25</sup> | 33 | *P. thiaminolyticus* | wound infection | none | Switzerland | 4 weeks amox-clav | cured |
| 2021<sup>19</sup> | 19 | *P. turicensis* | fracture nonunion | none | USA | 6 weeks ciprofloxacin | cured |
| 2023<sup>16</sup> | 57 | *P. silvae* | bacteremia | obesity | Italy | 2 weeks amox-clav +<br>doxycycline | cured |
| 2023<sup>26</sup> | 30 | *P. lactis* | brain abscess,<br>meningitis | IVDU | Bulgaria | vancomycin,<br>meropenem,<br>metronidazole (duration<br>unknown) | cured without<br>neurologic sequelae |

Abbreviations: ESRD: end-stage renal disease; CHF: congestive heart failure; IVDU: intravenous drug use; NTM: nontuberculous mycobacterium; ARDS: acute respiratory distress syndrome; NF1: neurofibromatosis type 1; tmp/smx: trimethoprim-sulfamethoxazole

One hundred and thirty-nine *Paenibacillus* infections in infants were reported in 9 papers. They included two separate Ugandan cohorts previously described by our group^4,5^ and 6 single case reports from throughout the United States (Table 2).^38–44^ In contrast to the infections occurring in adults, *Paenibacillus* infections in infants were caused by only 3 species: *P. thiaminolyticus* (112/139, 80%), *P. alvei* (2/139, 1%) and *P. dendritiformis* (2/139, 1%). An additional 25/139 (18%) were identified only to the genus level. All of the infants with *Paenibacillus* infection presented with a sepsis syndrome with or without clinical signs of meningitis, or with confirmed meningitis, often complicated by extensive cerebral destruction and hydrocephalus. Outcomes were commonly poor with 17% (24/139) mortality. Cystic encephalomalacia due to brain destruction was common in both Ugandan and American cases and 92/139 (66%) required surgical management of hydrocephalus following their infection. We have additionally received personal communications about 2 additional neonates with sepsis due to *Paenibacillus*, one from Minnesota and one from Maryland. One presented in extremis, was intubated and received extra corporeal support but succumbed to the illness within 2 days. Blood culture grew *Paenibacillus species*; CSF culture was not performed due to the infant’s critical illness. The other infant was unexpectedly delivered at home with unknown gestational age, thought to be term with a birthweight of 4082 grams. Maternal toxicology screen was positive for amphetamines, methamphetamines, and tetrahydrocannabinol. The infant developed tachypnea on day of life 1 and had an elevated C-reactive protein of 5.8 mg/L. Blood culture grew *Paenibacillus species*. Lumbar puncture was not performed due to caregiver decision. The infant remained clinically stable and was treated with 8 days of gentamicin; he was discharged in stable condition at 13 days old with no appreciable neurologic sequelae.

**Table 2.**
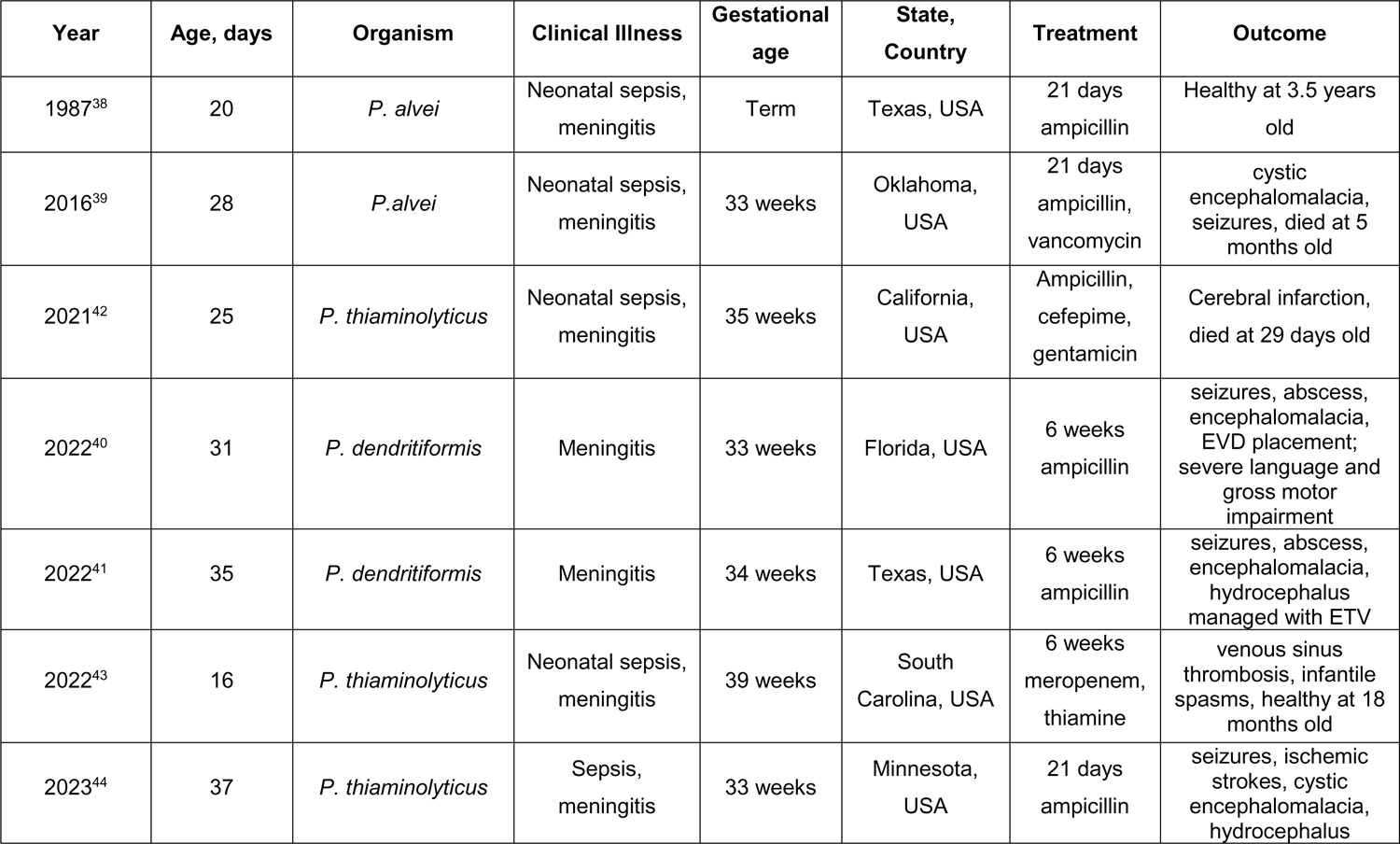
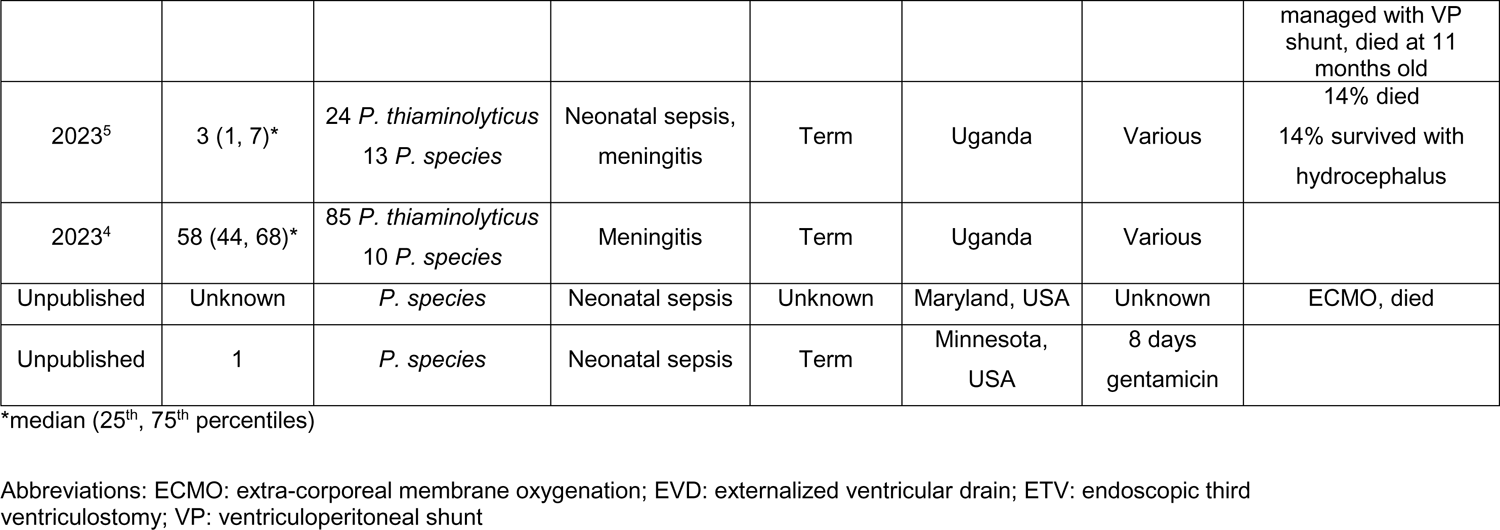
Reported infections due to *Paenibacillus species* in infants.

## Discussion

Based on rRNA homology and a distinct phenotypic cluster of Bacillus, the Paenibacillus genus was split off from Bacillus in 1993.^7^ Since then, it has generally been considered an environmental commensal with little concern for its role as a pathogen. There has been some interest in the genus for its ability to produce antibacterial compounds such as paenibacterin, a novel cyclic lipopeptide, which have activity against *Staphylococcus aureus*, *Pseudomonas aeruginosa,* and *Escherichia coli*.^45–47^ More recently, the possibility that *Paenibacillus* may be a useful probiotic with agricultural,^48–50^ aquacultural,^51,52^ and human^53^ applications has even been considered.

*Paenibacillus* growth on culture has sometimes been found to be due to contamination.^54^ We sought to compile all of the reports of human disease caused by *Paenibacillus species* to evaluate its role as a human pathogen. In doing so, we found that adult cases seem to be sporadic with no single clinical syndrome or species emerging as characteristic. Adult infections tended to occur in patients with comorbidities, indwelling hardware such as prosthetic joints or pacemakers, and were effectively treated with standard antibiotics in most cases. A single death was reported related to a *Paenibacillus* infection in an adult.^11^ This was in marked contrast to *Paenibacillus* infections in infants where most infections were due to *P. thiaminolyticus* and severe disease was common. Death occurred in 17% of cases and extensive destruction of the brain was common. Notably, there have been only two reported cases (5.4%) of *P. thiaminolyticus* in adults, with the bacteria isolated from the blood and the wounds of two patients in the United States and Switzerland, respectively.^13,25^ The current study is potentially limited by publication bias such that many clinical cases may not be reported in the literature. However, the pattern of reports suggests that *Paenibacillus* species’ cause a different pattern of illness in infants compared to older patients.

Interestingly, most cases in infants have been published within the last 3 years. This may be due to improvements in diagnostic technology: bacterial identification methods that use biochemical tests may misidentify *Paenibacillus* as a “*Bacillus* species” or otherwise be unable to resolve the bacterial identification down to the genus level. Now that more clinical laboratories are using matrix assisted laser desorption ionization-time of flight (MADLI-TOF) systems for pathogen identification, and the databases that underly this technology’s ability to identify pathogens has become more robust over time, specimens that were previously identified only as a Gram-positive rod, *Bacillus* or *Bacillus*-like bacteria are now properly being identified as *Paenibacillus* species. Another possible explanation is that changes in environmental ecology or other risk factors lead to more *Paenibacillus* infections than before. If the latter is true, then *P. thiaminolyticus*, and other *Paenibacillus* species, may represent emerging pathogens among infants.

Optimal treatment is not currently known, and attempts to use standard therapies in reported cases have frequently resulted in poor outcomes. Clinical trials to determine optimal antibiotic therapy of *Paenibacillus* neonatal sepsis and *Paenibacillus* central nervous system infections in infants are urgently needed. Because *P. thiaminolyticus* predominates as the cause of neonatal paenibacilliosis, adjunctive therapies such as thiamine supplementation may also be helpful.^43^ Thiamine deficiency has well-described neurologic sequelae (i.e. Wernicke’s encephalopathy) which may contribute to the severity of cerebral destruction in neonatal *Paenibacillus* infections.^55^ Supplementation has been proposed as a neuroprotective strategy for infants with hypoxic-ischemic encephalopathy.^56^ It is possible that thiamine supplementation could blunt the effects of local thiamine depletion that may in the brain when *P. thiaminolyticus* produces thiaminase as part of its typical metabolic pathways.^57^ Most of the infants for whom a treatment regimen was recorded received ampicillin with or without gentamicin. Ceftriaxone was also commonly administered. One American infant who had severe disease at the time of presentation with seizures, infarction of the thalamus and corona radiata, cystic encephalomalacia, venous sinus thrombosis and ultimately required an endoscopic third ventriculostomy for management of hydrocephalus, was found to have appropriate neurodevelopment at age 18 months after being treated with supplemental thiamine and 6 weeks of intravenous meropenem.^43^ It is not clear whether it was the meropenem, the thiamine or the combination of both that was instrumental in achieving this good outcome.

Importantly, resistance to penicillin, ampicillin, erythromycin and vancomycin was reported for both adult and infant *Paenibacillus* infections. Notably, Paenibacillus spp. can exhibit intrinsic resistance to critically important antibiotics due to the natural presence of multiple resistance genes, such as rifampin phosphotransferase (*rph*), chloramphenicol acetyltransferase (*cat*), chloramphenicol–florfenicol resistance (*cfr*), streptogramin B lyase (*vgb*), streptogramin A acetyltransferase (*vat*), and macrolide kinase (*mph*) genes.^58,59^ Antibiotic susceptibility results should be used to guide antimicrobial therapy. Ampicillin and vancomycin, antibiotics commonly used for Gram-positive neonatal infections, are not reliably effective for *Paenibacillus* infection.^5,15,22,24^ Additional caution should be exercised in using ampicillin because some *Paenibacillus* isolates have been shown to have beta-lactamase genes which could confer inducible resistance; the clinical relevance of these genes in human infections is not well established.^60^ Ceftriaxone seems to be susceptible in most human cases but has not been shown to achieve good clinical outcomes and resistance has been noted in environmental isolates.^5,61^ Meropenem has been successfully used in a few cases.^43^

Changes in environmental ecology, whether due to climate change or other factors, may increase human contact with *Paenibacillus* in the environment. Our work has suggested that neonates with paenibacilliosis are more like to have had non-sterile substances applied to their umbilical stumps.^5^ These neonates often also live in agrarian and herding communities with close contacts with livestock where use of antibiotics may impact antibiotic resistance in environmental reservoirs.^61,62^ An approach to neonatal paenibacilliosis that uses a One Health perspective to understanding the epidemiology, pathogenesis and treatment of this infection is likely to be more successful than one focused entirely on a single aspect of the disease.^63^

## Conclusion

Human infections due to *Paenibacillus* species’ have been sporadically reported in adults and have recently been described as causing a distinct clinical syndrome of sepsis, meningitis and multiple brain abscesses leading to cystic encephalomalacia and hydrocephalus in infants that we have termed “neonatal paenibacilliosis”. The high mortality and the severity of the sequelae secondary to neonatal paenibacilliosis warrant further and urgent investigation to better understand the mechanisms, risk factors and optimal treatments for this devastating infection. Given the evidence that *Paenibacillus* is an important neonatal pathogen, its use as a probiotic should be avoided until the safety of specific species has been carefully studied. Similarly, widespread application of the organism or its spores in agricultural applications should not be implemented until the pathophysiology of neonatal disease, route of neonatal exposure, prevention strategies and effective treatments are better understood. Neonatal paenibacilliosis requires further investigation in order to elucidate the role of *Paenibacillus* species as a possible emerging pathogen and to define optimal treatments to improve neurodevelopmental outcomes.

## Data Availability

All data produced in the present work are contained in the manuscript.

